# Population-based estimates of hepatitis E virus associated mortality in Bangladesh

**DOI:** 10.1101/2024.08.24.24312522

**Authors:** Repon C Paul, Heather F Gidding, Arifa Nazneen, Kajal C Banik, Shariful Amin Sumon, Kishor K Paul, Arifa Akram, M Salim Uzzaman, Alexandra Tejada-Strop, Saleem Kamili, Stephen P Luby, Andrew Hayen, Emily S Gurley

**Author notes:** Funding: The Centers for Disease Control and Prevention (CDC), United States.

## Abstract

**Background:** Hepatitis E virus (HEV) is endemic in many resource-poor countries. Despite an available vaccine, data on HEV-associated mortality are scarce, hindering informed decisions. This study aims to estimate the population-based rate of HEV-specific mortality in Bangladesh.

**Methods:** During December 2014-September 2017, we conducted surveillance in six tertiary hospitals in Bangladesh. Patients aged ≥14 years with acute jaundice were recruited, tested for IgM anti-HEV, and followed up post-discharge. A mortality survey in the hospital catchment areas identified deaths associated with acute jaundice, including maternal, stillbirths and neonatal deaths delivered by a mother with acute jaundice during pregnancy, confirmed by two independent physicians reviewing verbal autopsy data.

**Findings:** Out of 1,925 patients with acute jaundice identified and enrolled in the surveillance hospitals, 302 died, with 28 (9%) testing positive for IgM anti-HEV. In the hospital catchment areas, the team identified 587 jaundice-associated deaths, including 25 maternal deaths. Combining hospital-based surveillance and mortality survey data, the study estimated 986 (95% CI: 599-1338) HEV-associated deaths annually among individuals aged ≥14 years in Bangladesh, including 163 (95% CI: 57-395) maternal deaths. Additionally, 279 (95% CI: 101-664) stillbirths and 780 (95% CI: 365-1,297) neonatal deaths were attributed to HEV infection annually.

**Interpretation:** Prior Global Burden of Disease studies presented wildly varying modeling estimates of HEV-associated annual deaths, ranging from 50,000 in 2013 to 1,932 in 2019. This study is the first to directly measure population-based estimates of mortality in Bangladesh, which can be used to determine the cost-effectiveness of hepatitis E vaccination and other interventions.

## Introduction

Hepatitis E virus (HEV) causes acute liver infection; transmission of genotypes 1 and 2 occurs among humans through the fecal-oral route, while genotypes 3 and 4 are zoonotic and primarily infect humans through the consumption of undercooked meat^1,2^. In low- and middle-income countries, where fecal contamination of drinking water is common, genotypes 1 and 2 pose significant public health concerns^1,2^. In these settings, where HEV infection is a leading cause of acute viral hepatitis^1^, population-based studies estimating the disease burden are scarce due to inadequate surveillance systems and limited vital registration of cause-specific deaths.

Existing estimates of the burden of HEV infection rely on mathematical models, including the Global Burden of Disease effortwere built upon several strong assumptions^3,4^. These assumptions include predicting the proportion of people with antibodies against HEV who develop clinical disease, comparing HEV-associated mortality between HEV endemic low- and middle-income countries and high-income countries with effective vital registration, and extrapolating the likelihood of death among hospitalized hepatitis E patients to non-hospitalized severe cases. Notably, the global annual HEV-related death toll projections by these models have varied significantly, from 70,000 in 2005 to 1,932 in 2019^3-7^.

HEV infection is preventable through sanitation, adherence to safe food and water precautions, and potentially vaccination. Two hepatitis E vaccine candidates, following non-human primate experiments, progressed to clinical trials in humans^8,9^. One of these vaccines, HEV 239, was licensed in China in 2011 and in Pakistan in 2020 for use among residents aged ≥16 years^10^. A major barrier to recommending the vaccine in routine national vaccination programs in other countries, however, is the lack of confidence in the wide-ranging burden of disease estimates and a dearth of population-based measurements of HEV-associated mortality^11^.

During 2014-2017, the International Centre for Diarrhoeal Disease Research, Bangladesh (icddr,b) in collaboration with the Institute of Epidemiology, Disease Research and Control (IEDCR) of the Government of Bangladesh and the Centers for Disease Control and Prevention (CDC), United States conducted a hospital-based acute jaundice surveillance study in Bangladesh to identify patients admitted with acute jaundice^12^. However, estimating morality burden from hospital-based studies may not fully represent the general population due to healthcare access disparities in many low- and middle-income countries^13^. We conducted a mortality survey in the catchment areas of surveillance hospitals and calculated population-based estimates of HEV-specific mortality by combining data from the hospital-based surveillance and mortality survey.

## Methods

### Hospital-based acute jaundice surveillance

Detailed information on hospital-based acute jaundice surveillance is available elsewhere^12^. In summary, a study spanning December 2014 to September 2017 investigated acute jaundice in six randomly selected tertiary hospitals across five divisions in Bangladesh (Figure 1). Patients aged ≥14 years, exhibiting acute jaundice (defined as new onset of yellow sclera or skin during the past 3 months, persisting on admission day), were recruited. Blood samples from all recruited patients were tested for IgM anti-HEV using an enzyme-linked immunosorbent assay (ELISA) with a sensitivity of 98-100% and a specificity of 95-100% (Beijing Wantai Biologic Pharmacy Enterprise Co., Ltd, Beijing, China). Patients positive for IgM anti-HEV antibodies were considered to have acute HEV infection.

**Figure 1:**
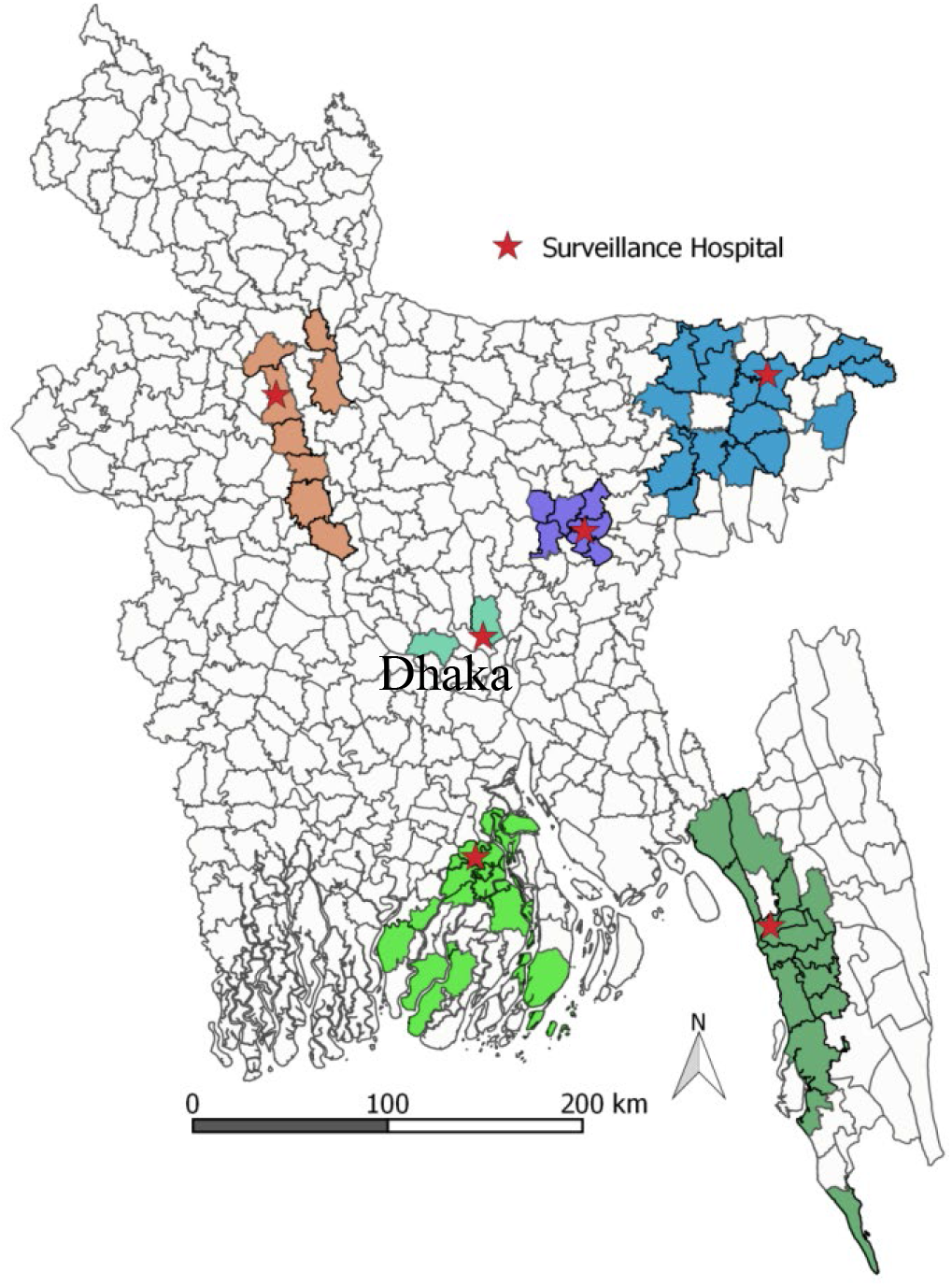
Map of Bangladesh showing the acute jaundice surveillance sites and the selected areas (sub-districts) for mortality survey, December 2014-September 2017.

Enrolled patients were followed during their hospitalization and for 3 months post-discharge to ascertain vital status and pregnancy outcomes for pregnant women. However, post-discharge follow-up was not possible for patients admitted after June, 2017, as surveillance ended on September 30, 2017.

### Mortality survey in hospital catchment areas

#### Selection of hospital catchment areas

The study team reviewed the hospital records of jaundice patients admitted from January to June 2014 to identify the primary catchment areas of each surveillance hospital, defined as the sub-districts where 75% of the admitted jaundice patients resided. Applying this criterion, we selected 63 sub-districts accorss 18 districts (out of 64) in Bangladesh. Sub-districts in Bangladesh consist of a median of 15 unions (IQR: 7-20 unions), which are the smallest administrative units in Bangladesh with an average population of 26,000 people^14^.

#### Selection of survey clusters

For rare events, such as maternal deaths, a large sample size is required to estimate death rates with acceptable precision^15^. Therefore, we assumed that our study, designed to estimate maternal mortality associated with HEV infection with acceptable precision, would also provide sufficient power for estimating mortality in adults, neonatal deaths, and stillbirths associated with HEV infection. The sample size for estimating maternal mortality was determined using the formula for estimating proportions^16^, assuming that 10% of maternal deaths are associated with HEV infection^17^. With a precision of 5%, a 95% confidence level, and a design effect of 2.0, we calculated a required sample size of 277 maternal deaths in the surveillance hospitals’ catchment areas. To achieve this size, we conducted the survey in 91 unions, collecting mortality data for the prior three years. Assuming a maternal mortality ratio of 194 per 100,000 live births^18^ and the crude live birth rate of 22.2 per 1,000 population^19^, we randomly selected 91 unions from the defined hospital catchment areas using a probability proportional to size sampling approach^20^.

#### Mortality Survey

Between November 2014 and February 2017, a mortality survey was undertaken across 91 unions to identify deaths associated with jaundice. The survey also identified stillborn babies and live-born babies who died during neonatal period delivered by mothers with jaundice during pregnancy, and all-cause maternal deaths (Figure 1). Jaundice was defined as having new onset of either yellow sclera or skin during the illness prior to death. A neonatal death was defined as occuring within the first 28 days of life, while a stillbirth was defined as a baby delivered by a mother after 28 weeks of gestation with no signs of life^21^. Maternal death was defined as the death of a woman while pregnant or within 42 days of pregnancy termination, excluding unintentional injuries or unrelated incidental causes^21^.

In urban communities, we conducted a house-to-house survey to identify deaths, whereas in rural communities, we used a low-cost community knowledge approach (further detailed in^22^). Out of 91 unions, 25 (27%) were urban, closely matching the urban population percentage (23%) in the 2011 Bangladesh Population Census^23^. For identified jaundice-associated deaths and all-cause maternal deaths, a verbal autopsy questionnaire was administered to collect detailed information on the signs, symptoms and duration of illness preceding death to determine the underlying and immediate cause of death. Two physicians independently assessed VA questionnaires, assigning ICD-10 cause of death codes. Discordant cases prompted discussion for consensus, with a third physician intervening if disagreement persisted.

### Data analysis

We calculated the HEV-associated mortality rate for individuals aged ≥14 years, maternal mortality ratio, and stillbirth and neonatal mortality rate for all catchment areas combined. The population in 2014 served as our mid-year reference, considering our data collection spanned from November 2014 and February 2017 (about 60% collected in 2015) and we collected mortality data for the prior three years. For rural unions, we projected the population for 2014 based on the 2011 Bangladesh census, considering an annual growth rate of 1.37% (Appendix 1)^24^. For the urban unions, we counted the household population at the time of mortality survey. We projected the population aged ≥14 years, assuming 67.7% of total population in 2014 were aged ≥14 years^24^. The number of live births in 2014 was estimated assuming a crude birth rate of 22.2 per 1,000 population as per the Bangladesh Demographic and Health Survey, 2014^19^.

Although we collected information on both chronic and acute jaundice associated deaths in the mortality survey, for incidence calculation only deaths associated with acute jaundice were considered, defined as new onset of yellow eyes or skin within the 3 months prior to death.

Because we used the same case definition of acute jaundice in hospitals and hospital catchment areas, we assumed that the illness in people who died in the catchment areas was similar to those who died in the study hospitals. To calculate HEV associated mortality in the hospital catchment areas, we multiplied the estimate for jaundice-associated mortality in the hospital catchment areas by the proportion of HEV infections among acute jaundice deaths in the surveillance hospitals.

Recognizing HEV’s additional risk during pregnancy^1,2^ and its varying age-specific infection rates and severity^25^, we separately projected HEV-associated deaths among pregnant women and non-pregnant persons aged ≥14 years (14-44 years, 45-59 years, and ≥ 60 years). The projected deaths in both groups were aggregated to estimate the total number of HEV-associated deaths in the hospital catchment areas. We used the following equation to estimate mortality associated with HEV in the hospital catchment areas:

HEV associated mortality for the population aged ≥14 years =

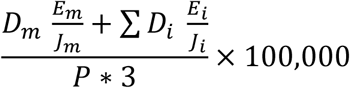

where,

*P* = Estimated population aged ≥14 years in the study areas in 2014

*D*_*m*_ =Number of maternal deaths associated with acute jaundice identified in the three-year period in the study areas

*J*_*m*_ = Number of maternal deaths associated with acute jaundice in the surveillance hospitals

*E*_*m*_ = Number of laboratory-confirmed HEV cases among maternal deaths associated with acute jaundice in the surveillance hospitals

*D*_*i*_ = Number of age-specific non-maternal deaths associated with acute jaundice identified in the three-year period in the study areas

*J*_*i*_ = Number of age-specific non-maternal deaths associated with acute jaundice in the surveillance hospitals

*E*_*i*_ = Number of age-specific laboratory-confirmed HEV cases among non-maternal deaths associated with acute jaundice in the surveillance hospitals

Similarly, the following formulas were used to calculate HEV associated maternal mortality, neonatal mortality and stillbirth:

Maternal mortality ratio associated with 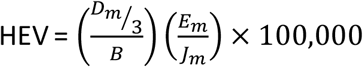

Stillbirth rate associated with 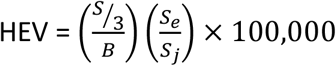

Neonatal mortality rate associated with 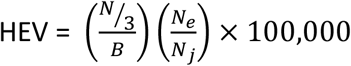 where, *D*_*m*_, *J*_*m*_ and *E*_*m*_ are defined above and:

*B* = Estimated live births in the study areas in 2014

*S* = Number of stillbirths delivered by a mother with acute jaundice during pregnancy identified in the three-year period in the study areas

*S*_*j*_ = Number of stillbirths delivered by a mother with acute jaundice during pregnancy in the surveillance hospitals

*S*_*e*_ = Number of stillbirths delivered by a mother with acute HEV during pregnancy in the surveillance hospitals

*N* = Number of neonatal deaths born to a mother with acute jaundice during pregnancy identified in the three-year period in the study areas

*N*_*j*_ = Number of neonatal deaths born to a mother with acute jaundice during pregnancy in the surveillance hospitals

*N*_*e*_ *=* Number of neonatal deaths born to a mother with acute HEV during pregnancy in the surveillance hospitals

We used non-parametric bootstrapping^26^ to estimate the 95% confidence intervals for hepatitis E associated mortality for the population aged ≥14 years. The 95% confidence intervals for maternal mortality, stillbirth and neonatal mortality associated with HEV infection were calculated by multiplying the lower and upper bounds of the jaundice associated mortality in the hospital catchment areas by the respective lower and upper bounds of the proportion of HEV infection among acute jaundice deaths in the study hospitals. The 95% confidence interval of the proportion of hepatitis E among acute jaundice deaths in the study hospitals was calculated by using the Wilson method for a binomial distribution^27^, and the 95% confidence interval for jaundice associated deaths in the hospital catchment areas was calculated using the confidence interval for the mean of a Poisson distribution. We performed a sensitivity analysis to check how the estimates of HEV-associated mortality vary when restricted to a two-year and one-year recall period.

### Population mortality estimates

The hepatitis E-associated mortality estimates generated from the study hospitals and the mortality surveys in the hospital catchment areas were considered representative of the hospital catchment areas, and with surveillance hospitals covering 5 out of 7 administrative divisions, we expected that results could be extrapolated to the whole country (Figure 1). The rural to urban population ratio of our study matched the national ratio, which was important because urbanicity is a known risk factor for HEV infection^23^. We, therefore, used our morality estimates to extrapolate the total number of HEV-associated deaths among the population aged ≥14 years, maternal deaths associated with HEV, stillborn babies and live born babies who died during the neonatal period delivered by mothers with acute HEV infection in pregnancy in 2014 in Bangladesh.

### Ethical approval and consent

The study protocol was reviewed and approved by the institutional review board of the icddr,b (Protocol # PR-14060). In the surveillance hospitals, written informed consent was obtained from patients over 17 years. For patients aged 14-17 years, assent was taken from the patients and written consent from their parents. Written consent was sought from their guardians or accompanying attendants if the patients were unable to provide consent themselves because of illness. In the mortality survey in the hospital catchment areas, written consent was obtained from a family member of the deceased directly involved in caregiving during the illness period.

## Results

### Deaths among hospitalized acute jaundice patients

In the six study hospitals, 1,925 acute jaundice patients were identified during December 2014 to September 2017; all patients consented for enrolment. Of them, 1,765 patients (92%; including 173 pregnant women) were followed-up during hospitalization and three months post-discharge. Among these patients, 302 died (17%), including 27 maternal deaths, and 28 of these deaths (9%; 95% CI: 6%-13%) tested positive for IgM anti-HEV. Among the maternal deaths, 8 cases (30%; 95% CI: 16%-49%) were positive for IgM anti-HEV (Table 1).

**Table 1:** Hepatitis E cases among patients with acute jaundice who died during hospitalization or within three months of hospital discharge in six tertiary hospitals in Bangladesh, December 2014–September 2017.

| Characteristics | Patients with acute jaundice who died* (N=302) | IgM Anti-HEV Positive (N=28) |  |
| --- | --- | --- | --- |
|  |  | n | Percent (95% CI) |
| Maternal deaths | 27 | 8 | 29.6 (15.8-48.5) |
| Non-maternal deaths | 275 | 20 | 7.2 (4.8-11.0) |
| Sex |  |  |  |
| Male | 178 | 13 | 7.3 (4.3-12.1) |
| Female | 97 | 7 | 7.2 (3.5-14.2) |
| Age group |  |  |  |
| 14-44 years | 96 | 14 | 14.6 (8.9-23.6) |
| 45-59 years | 86 | 3 | 3.5 (1.2-9.8) |
| 60+ years | 93 | 3 | 3.2 (1.1-9.1) |

\* Defined as new onset of either yellow eyes or skin during the past 3 months, continuing on the day of admission.

Among the 173 pregnant women followed, 125 (72%) had live births, 27 (16%) experienced stillbirths, 11 (6%) had miscarriages, and 10 (6%) women died before the pregnancy outcome. Of the enrolled pregnant women, 66 (38%) tested positive for IgM anti-HEV. Six (22%; 95% CI: 11%-41%) of the 27 stillbirths were delivered by mothers with IgM anti-HEV positivity. Among the live births, 15 (12%) died within 28 days of their birth (neonatal deaths). Ten (67%; 95% CI: 42%-85%) of the 15 neonatal deaths were born to mothers who were positive for IgM anti-HEV.

### Acute jaundice deaths in the hospital catchment areas

In the catchment areas of study hospitals, the team identified 33,794 deaths among persons aged ≥14 years who died in the three years prior to the date of survey (Table 2). Among these, 277 (0.8%) were maternal deaths. Of the maternal deaths, 25 (9%) had acute jaundice, and of the non-maternal deaths, 587 (1.7%) had acute jaundice prior to death. There were 57 stillbirths and 53 neonatal deaths delivered by mothers with acute jaundice during pregnancy identified.

**Table 2:** Deaths associated with acute jaundice in the catchment areas of the six surveillance hospitals, Bangladesh during the three-year period prior to the survey (Survey period: November 2014–February 2017)

| Characteristics | Deaths |  | Total |
| --- | --- | --- | --- |
|  | aged ≥ 14 years |  |  |
|  | Maternal deaths | Non-maternal deaths |  |
| Deaths identified in the study areas* | 277 | 33,517 | 33,794 |
| Deaths with jaundice<sup>†</sup> prior to death | 28 (10.1%) | 778 (2.3%) | 806 (2.4%) |
|  | [95% CI: 7.0%-14.2%] | [95% CI: 2.2%-2.5%] | [95% CI: 2.2%-2.6%] |
| Deaths with acute jaundice<sup>‡</sup> prior to death | 25 (9.0%) | 587 (1.7%) | 612 (1.8%) |
|  | [95% CI: 6.1%-12.9%] | [95% CI: 1.6%-1.9%] | [95% CI: 1.7%-1.9%] |

\* There were two more maternal deaths for whom their caregivers refused to participate in this study.
<sup>†</sup> Defined as having yellow eyes or skin prior to death
<sup>‡</sup> Defined as new onset of either yellow eyes or skin during the 3 months prior to death

### Demographic characteristics of acute jaundice deaths

#### Surveillance hospitals

Among non-maternal acute jaundice deaths, about two-thirds were males, and one-third were aged ≥60 years (Table 3). Eighty-two percent resided in rural areas, and 40% did no formal education. In most cases (72%), jaundice duration before hospitalization was less than one month, and over three-quarters of the deaths occurred at home post-discharge.

**Table 3:** Demographic characteristics of enrolled patients with acute jaundice in the study hospitals who died during hospitalization or within three months of hospital discharge and identified deaths associated with acute jaundice in the hospital catchment areas during the three-year period prior to the survey.

| Characteristics | Deaths* in hospitals |  |  | Deaths* in hospital catchment areas |  |  | P-value† |
| --- | --- | --- | --- | --- | --- | --- | --- |
|  | Maternal | Non-maternal | Total | Maternal | Non-maternal | Total |  |
|  | N=27<br>n (%) | N=275<br>n (%) | N=302<br>n (%) | N=25<br>n (%) | N=587<br>n (%) | N=612<br>n (%) |  |
| Sex |  |  |  |  |  |  | 0.288 |
| Male | - | 178 (65) | 178 (59) | - | 383 (65) | 383 (63) |  |
| Female | 27 (100) | 97 (35) | 124 (41) | 25 (100) | 204 (35) | 229 (37) |  |
| Age in years |  |  |  |  |  |  | <0.001 |
| 14-44 | 27 (100) | 96 (35) | 123 (41) | 25 (100) | 158 (27) | 183 (30) |  |
| 45-59 | 0 | 86 (31) | 86 (29) | 0 | 160 (27) | 160 (26) |  |
| ≥ 60 | 0 | 93 (34) | 93 (31) | 0 | 269 (46) | 269 (44) |  |
| Area of residence |  |  |  |  |  |  | 0.036 |
| Rural | 17 (63) | 232 (84) | 249 (82) | 19 (76) | 517 (88) | 536 (88) |  |
| Urban | 10 (37) | 43 (16) | 53 (18) | 6 (24) | 70 (12) | 76 (12) |  |
| Education |  |  |  |  |  |  | <0.001 |
| None | 4 (15) | 118 (43) | 122 (40) | 5 (20) | 306 (52) | 311 (51) |  |
| Class 1-4 | 8 (30) | 72 (26) | 80 (27) | 3 (12) | 61 (10) | 64 (11) |  |
| Class 5-9 | 9 (33) | 55 (20) | 64 (21) | 16 (64) | 160 (27) | 176 (29) |  |
| Class 10 or more | 6 (22) | 30 (11) | 36 (12) | 1 (4) | 49 (8) | 50 (8) |  |
| Unknown | 0 | 0 | 0 | 0 | 11 (2) | 11 (2) |  |
| Duration of jaundice at the time of death |  |  |  |  |  |  | 0.002 |
| ≤ 1 month | 21 (78) | 197 (72) | 218 (72) | 16 (64) | 356 (61) | 372 (61) |  |
| 1< and ≤ 2 months | 5 (19) | 49 (18) | 54 (18) | 1 (4) | 138 (24) | 139 (23) |  |
| > 2 and ≤ 3 months | 1 (4) | 29 (11) | 30 (10) | 8 (32) | 93 (16) | 101 (17) |  |
| Sought care from a qualified health care provider‡ |  |  |  |  |  |  |  |
| First visit | - | - | - | 15 (60) | 343 | 358 (59) |  |
| First or second visit | - | - | - | 20 (80) | 481 | 501 (82) |  |
| Any visit | - | - | - | 21 (84) | 544 | 565 (92) |  |
| Place of death |  |  |  |  |  |  | 0.103 |
| Health facility | 14 (52) | 51 (19) | 65 (22) | 15 (60) | 85 | 100 (16) |  |
| Home | 13 (48) | 224 (81) | 237 (78) | 7 (28) | 480 | 487 (80) |  |
| In-transit |  |  |  | 3 (12) | 22 | 25 (4) |  |

\* Among persons aged ≥ 14 years
† Chi-squared test comparing total deaths in hospitals and hospital catchment areas by patients' characteristics
‡ Health facility/qualified medical doctor/paramedic/nurse

#### Hospital catchment areas

The sex ratio of acute jaundice deaths identified in the hospital catchment areas was similar to those in the surveillance hospitals (p=0.288) (Table 3). However, the patients who died in the hospital catchment areas were older than the patients who died after enrolment in the hospital surveillance study (p <0.001).

### Population-based estimate of HEV associated mortality

In the 91 unions selected from the catchment areas of six study hospitals, the projected population aged ≥14 years was approximately 1.6 million, and the projected number of live births was 52,326 in 2014 (Appendix 1). We estimated HEV-associated mortality as 0.9 (95% CI: 0.6-1.3) per 100,000 population aged ≥14 years (Table 4). The maternal mortality ratio associated with HEV was estimated as 4.7 (95% CI: 1.6-11.4) per 100,000 live births. We estimated the HEV associated stillbirth rate as 8.1 (95% CI: 2.9-19.2) and neonatal mortality rate as 22.5 (95% CI: 10.5-34.5) per 100,000 live births. We extrapolated these estimates to the whole population of Bangladesh and estimated that in 2014 there were 986 (95% CI: 599-1338) HEV-associated deaths among the population aged ≥14 years including 163 (95% CI: 57-395) maternal deaths. Additionally, we estimated 279 (95% CI: 101-664) stillbirths and 780 (95% CI: 365-1,297) neonatal deaths associated with HEV infection in 2014 in Bangladesh.

**Table 4:**
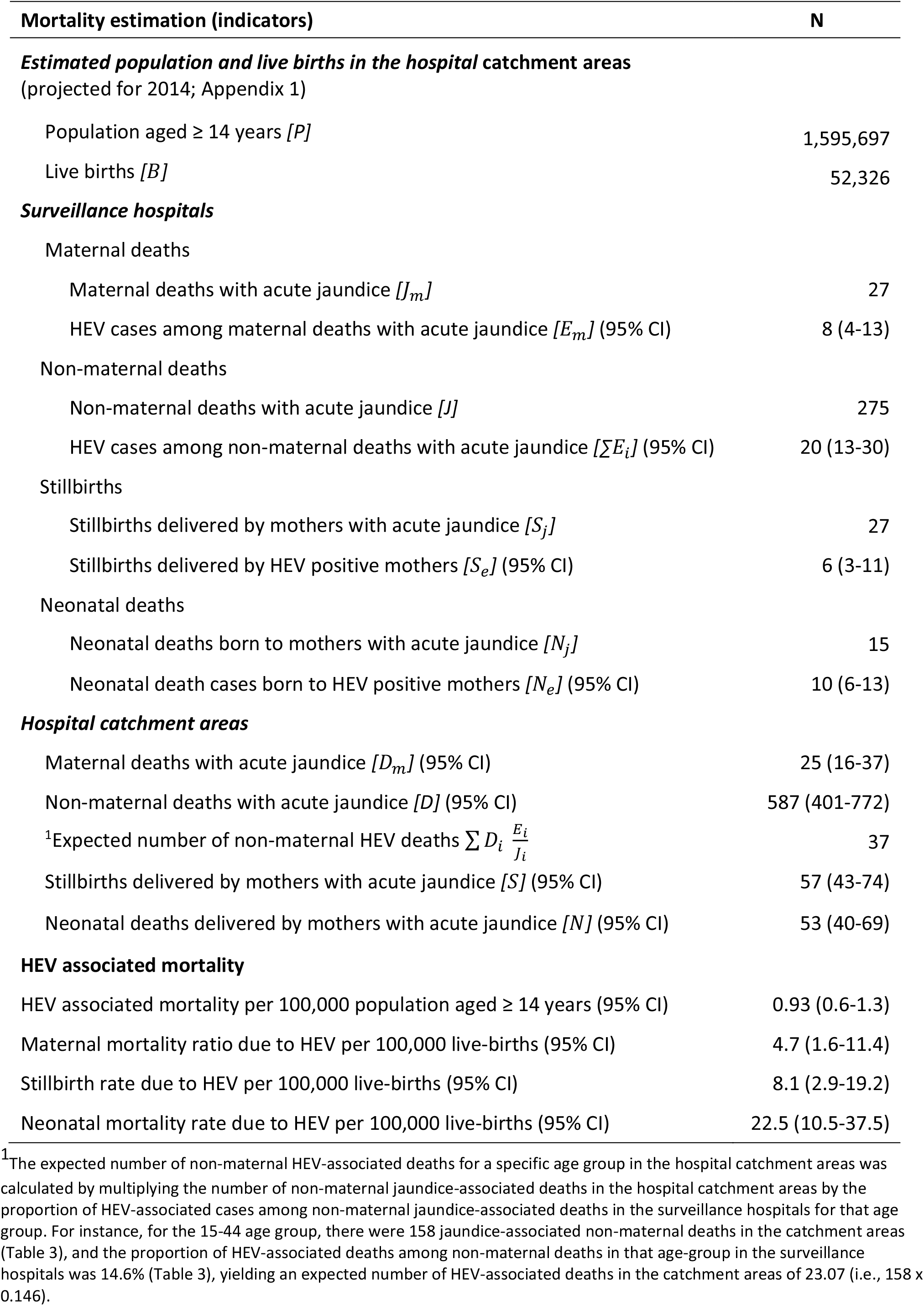
Estimation of HEV associated mortality in the catchment areas of six acute jaundice surveillance hospitals in Bangladesh, 2014.

In the sensitivity analysis for a two-year recall period, the estimated annual number of HEV-associated deaths were 1,114 (95% CI: 634-1,584) for population aged ≥14 years (including 186 (95% CI: 60-476) maternal deaths), 375 (95% CI: 133-904) stillbirths and 780 (95% CI: 337-1,367) neonatal deaths (Appendix 2). For one year recall period, the estimated number of HEV-associated annual deaths were 1,101 (95% CI: 422-1,901) for population aged ≥14 years (including 177 (95% CI: 43-548) maternal deaths), 412 (95% CI: 131-1092) stillbirths and 927 (95% CI: 359-1802) neonatal deaths.

## Discussion

This study provides the first measured estimate of HEV associated mortality in Bangladesh by combining data from a mortality survey covering a population of over 2.3 million and hospital-based acute jaundice surveillance in six hospitals across the country. In the absence of any reported HEV outbreak in Bangladesh during 2013-2015, we estimated that HEV infection accounted for 1766 (95% CI: 964-2635) deaths annually, including pregnant women and neonates. Additionally, we estimated 279 stillbirths per year, which are excluded from global burden of disease estimates^7^, despite their significant physical and psychological toll. Preventing HEV infections during pregnancy could prevent an estimated 1,222 maternal and neonatal deaths and stillbirths annually. This population-based estimate of HEV mortality in Bangladesh can inform the cost-effectiveness of HEV vaccination and other control measures to help prioritise public health interventions.

The Global Burden of Disease (GBD) studies provide estimates of global and country-level disease burdens, including hepatitis E, by employing models based on available data from vital registration, surveillance, and verbal autopsy^7^. Most recently, the 2019 GBD study estimated that there were 1,932 HEV-associated deaths globally in 2019^7^, which appears to be a significant underestimate as we identified approximately this many deaths in Bangladesh alone. The HEV mortality estimate by GBD studies appears to be sharp reduction, from 70,000 in 2005, 50,000 in 2013, 26,100 in 2016 and 14,700 in 2017^3-6^. This sudden decrease is likely due to methodological changes in calculation and the use of different data sources across various GBD studies. For instance, GBD studies transitioned from a single cause-of-death ensemble model (CODEm) to a two-model hybrid approach in 2013, incorporating a global CODEm model and a CODEm model limited to data-rich countries^3,4^,7. Moreover, GBD 2019 considered location- and year-specific factors for virus-specific case fatality rates^7^.

The 2013 global burden of disease study estimated 38,738 viral hepatitis deaths (25 deaths/100,000 population) in Bangladesh, with 12.7% attributed to HEV, totalling 4,920 HEV-associated deaths^4^. However, subsequent GBD studies conducted in 2017 and 2019 reported significantly lower figures of 227 and 111 HEV-associated deaths, respectively^6,7^. These variations and model uncertainties have left decision-makers hesitant on efficient vaccine implementation.

We note a number of study limitations. First, in the hospital catchment areas, the signs of jaundice among deaths were not verified by medical professionals; instead, caregivers reported symptoms of jaundice in decedents who had passed away within 3 years of the survey. Although our trained field team explained jaundice symptoms, and laypersons can identify eye yellowing when pronounced, misclassification may have occurred. This limitation in common in mortality surveys using the verbal autopsy method. Second, conducting interviews with caregivers promptly is crucial for accurate cause-of-death identification, as delays between death and the survey may introduce bias in reported symptoms^28^. Our sensitivity analysis showed slightly lower number of identified jaundice-associated deaths in the third year preceding the survey compared to the first two years. While uncertain if this reflects a true change in burden or random fluctuation, we provided separate estimations for HEV-associated deaths based on the one and two years recall periods, illustrating how estimates vary when restricting recall bias (Appendix 2). Third, while the Wantai IgM anti-HEV ELISA test is widely accepted as highly sensitive for diagnosing acute HEV infections, a study by Huang et al. observed that about 3% of acute viral hepatitis cases were negative for IgM anti-HEV but had a 4-fold rise in IgG anti-HEV in their convalescent sera, indicating potential underestimation in our prevalence estimates due to the test’s sensitivity^29^. Fourth, our assessment of HEV mortality in Bangladesh did not account for miscarriages, which is significant among pregnant women with HEV infection^30^. Finally, this study presents the HEV burden in Bangladesh based on data six to nine years old, which may not precisely reflect the current situation. Nonetheless, there are no other measured estimates of HEV mortality in Bangladesh, so our estimates remain useful. Since there have been no targeted strategies to reduce HEV transmission since this study, it is unlikely that there has been a significant change in HEV burden in Bangladesh over this period.

This study provides a dependable estimate of HEV associated mortality in Bangladesh in the absence of reliable data on HEV associated mortality in Bangladesh or in any region of the world^11^. Data from this study highlight the substantial underestimates of HEV deaths globally, based on the latest global burden of disease estimates published in 2020^7^, suggesting that these estimates should not be used for making decisions about global recommendations or policies.

Our data from Bangladesh can help determine the cost-effectiveness of vaccination against HEV and other interventions, including improvement of drinking water quality, for national policymakers, and our methods can be used by other countries requiring accurate HEV-associated mortality estimates.

Disclaimer: The findings and conclusions in this report are those of the authors and do not necessarily reflect the official position of the Centers for Disease Control and Prevention, or the authors’ affiliated institutions.

## Data Availability

All data produced in the present study are available upon reasonable request to the authors

