## Supplementary Tables for "Population-based estimates of hepatitis E virus associated mortality in Bangladesh"

### Appendices

#### Appendix 1: Projected population and live-births in 2014 in the catchment areas of the six acute jaundice surveillance hospitals, Bangladesh

| | | | Population projection[2] | | | | |
|---|---|---|---|---|---|---|---|
| **Surveillance hospitals** | **No. of unions** | **Census population[1]** | **2012** | **2013** | **2014** | **Projected population ≥ 14 years[4]** | **Projected live-birth[5]** |
| Bogra | | | | | | | |
| Rural | 12 | 427,635 | 433,494 | 439,432 | 445,453 | | |
| Urban[3] | 2 | 34,414 | - | | - | | |
| ***Total*** | ***14*** | | | | ***479,867*** | ***324,870*** | ***10,653*** |
| Barisal | | | | | | | |
| Rural | 11 | 214,544 | 217,483 | 220,463 | 223,483 | | |
| Urban[3] | 4 | 49,278 | - | | - | | |
| ***Total*** | ***15*** | | | | ***272,761*** | ***184,659*** | ***6,055*** |
| Kishoregonj | | | | | | | |
| Rural | 6 | 153,425 | 155,527 | 157,658 | 159,818 | | |
| Urban[3] | 1 | 3,645 | - | | - | | |
| ***Total*** | ***7*** | | | | ***163,463*** | ***110,664*** | ***3,629*** |
| Chittagong | | | | | | | |
| Rural | 19 | 548,491 | 556,005 | 563,623 | 571,344 | | |
| Urban[3] | 6 | 101,065 | - | | - | | |
| ***Total*** | ***25*** | | | | ***672,409*** | ***455,221*** | ***14,927*** |
| Sylhet | | | | | | | |
| Rural | 15 | 393,194 | 398,581 | 404,041 | 409,577 | | |
| Urban[3] | 4 | 47,585 | - | | - | | |
| ***Total*** | ***19*** | | | | ***457,162*** | ***309,498*** | ***10,149*** |
| Mitford, Dhaka | | | | | | | |
| Rural | 3 | 107,617 | 109,091 | 110,586 | 112,101 | | |
| Urban[3] | 8 | 199,249 | - | | - | | |
| ***Total*** | ***11*** | | | | ***311,350*** | ***210,784*** | ***6,912*** |
| All | | | | | | | |
| Rural | 66 | 1,844,906 | 1,870,181 | 1,895,803 | 1,921,775 | *1301,042* | *42,663* |
| Urban[3] | 25 | 435,236 | - | | 435,236 | *294,655* | *9,662* |
| ***Total*** | ***91*** | ***2,280,142*** | | | ***2,357,011*** | ***1,595,697*** | ***52,326*** |

[1] Population census in Bangladesh was conducted in 2011

[2] Projected population considering 1.37% growth rate

[3] Population in urban areas was counted through house-to-house visits in the mortality survey

[4] Projected considering 67.7% of population are ≥ 14 years

[5] Live-births are projected considering crude birth rate as 22.2/1000 population

#### Appendix 2: Estimation of HEV associated mortality in the catchment areas of six acute jaundice surveillance hospitals by recall period, 2014 (Survey period: November 2014–February 2017

| Mortality estimation (indicators) | Recall period | | |
|---|---|---|---|
| | **One year** | **Two years** | **Three years** |
| ***Estimated population and live births in the hospital* catchment areas** (projected for 2014; Appendix 1) | | | |
| Population aged ≥ 14 years *[P]* | 1,595,697 | 1,595,697 | 1,595,697 |
| Live births $[B]$ | 52,326 | 52,326 | 52,326 |
| ***Surveillance hospitals*** | | | |
| Maternal deaths | | | |
| Maternal deaths with acute jaundice $[J_m]$ | 27 | 27 | 27 |
| HEV cases among maternal deaths with acute jaundice $[E_m]$ (95% CI) | 8 (4-13) | 8 (4-13) | 8 (4-13) |
| Non-maternal deaths | | | |
| Non-maternal deaths with acute jaundice $[\sum J_i]$ | 275 | 275 | 275 |
| HEV cases among non-maternal deaths with acute jaundice $[\sum E_i]$ (95% CI) | 20 (13-30) | 20 (13-30) | 20 (13-30) |
| Stillbirths | | | |
| Stillbirths delivered by mothers with acute jaundice $[S_j]$ | 27 | 27 | 27 |
| Stillbirths delivered by HEV positive mothers $[S_e]$ (95% CI) | 6 (3-11) | 6 (3-11) | 6 (3-11) |
| Neonatal deaths | | | |
| Neonatal deaths born to mothers with acute jaundice $[N_j]$ | 15 | 15 | 15 |
| Neonatal death cases born to HEV positive mothers $[N_e]$ (95% CI) | 10 (6-13) | 10 (6-13) | 10 (6-13) |

| Mortality estimation (indicators) | Recall period | | |
|---|---|---|---|
| | One year | Two years | Three years |
| ***Hospital catchment areas*** | | | |
| Maternal deaths with acute jaundice [$D_m$] (95% CI) | 9 (4-17) | 19 (11-30) | 25 (16-37) |
| Non-maternal deaths with acute jaundice [$\sum D_i$] (95% CI) | 214 (186-245) | 437 (401-480 | 587 (401-772) |
| Stillbirths delivered by mothers with acute jaundice [$S$] (95% CI) | 28 (19-40) | 51 (38-67) | 57 (43-74) |
| Neonatal deaths born to mothers with acute jaundice [$N$] (95% CI) | 21 (13-32) | 35 (24-49) | 53 (40-69) |
| **HEV associated mortality** | | | |
| HEV associated mortality per 100,000 population aged ≥ 14 years (95% CI) | 1.0 (0.4-1.8) | 1.1 (0.6-1.5) | 0.93 (0.6-1.3) |
| Maternal mortality ratio due to HEV per 100,000 live-births (95% CI) | 5.1 (1.2-15.8) | 5.3 (1.7-13.7) | 4.7 (1.6-11.4) |
| Stillbirth rate due to HEV per 100,000 live-births (95% CI) | 11.9 (3.8-31.5) | 10.8 (3.8-26.1) | 8.1 (2.9-19.2) |
| Neonatal mortality rate due to HEV per 100,000 live-births (95% CI) | 26.7 (10.3-52.0) | 22.3 (9.7-39.4) | 22.5 (10.5-37.5) |
| **Estimated HEV associated annual deaths in Bangladesh** | | | |
| Deaths among population aged ≥ 14 years | 1102 (422-1901) | 1114 (634-1584) | 986 (599-1338) |
| Maternal deaths | 177 (43-548) | 186 (60-476) | 163 (57-395) |
| Stillbirths | 412 (131-1092) | 375 (133-904) | 279 (101-664) |
| Neonatal deaths | 927 (359-1802) | 780 (337-1367) | 780 (365-1298) |
